# Clinical application of interactive monitoring of indicators of health in professional dancers

**DOI:** 10.1101/2021.09.25.21263895

**Authors:** Nicola Keay, Martin Lanfear, Gavin Francis

**Affiliations:** Department of Sport and Exercise Sciences, Durham University, United Kingdom; Head of Performance Medicine, Scottish Ballet; Science4Performance, London, United Kingdom

## Abstract

**Objectives:** The purpose of this study was to assess the effectiveness of interactive monitoring of professional dancer health with a variety of subjective and objective monitoring methods and delivering swift personalised clinical advice.

**Methods:** Dancers from a ballet company completed a published, online dance-specific health questionnaire. Over the study period, dancers recorded wellbeing and training metrics, with menstrual cycle tracking and capillary blood testing for the recognised indicators of low energy availability. At regular, virtual clinical discussions with each dancer, findings were discussed and personalised advice given.

**Results:** Twenty dancers participated in the study (mean age 26.2 years, SD 3.7), comprising 14 females (mean age 25.5 years, SD 3.7) and 6 males (mean age 27.7 years, SD 2.4). Ten of the female and all the male dancers recorded positive scores on the dance health questionnaire, suggesting a low risk of relative energy deficiency in sport (RED-S). Two female dancers were taking hormonal contraception. Apart from one, all female dancers not on hormonal contraception reported current eumenorrhoeic status. Blood testing confirmed the low risk of insufficient energy availability across the group, apart from female dancers with negative questionnaire scores. The initiative of monitoring menstrual cycles and on demand virtual clinical support was well received by dancers, healthcare and artistic staff.

**Conclusions:** Multimodal monitoring facilitated delivery of prompt personalised clinical medical feedback specific for dance. This interactive strategy permitted the early identification and swift management of emergent clinical issues. Dancers highly rated the new monitoring modalities and opportunity to discuss health and well-being, in confidence, with a doctor conversant in dance.

**Summary boxes:** *What are the new findings?:*

- Monitoring professional dancers with a variety of interactive methods: dance specific questionnaire; online tracking of menstrual cycle for female dancers; wellbeing and training load; blood testing and personal online meetings - facilitates comprehensive, personalised support for dancer health.
- Overall, the dancer participants were healthy both physically and mentally. This was demonstrated by the positive scores derived from the dancer health questionnaire. The healthy status of the majority of the dancers was confirmed with online health tracking and objective blood test results.
- All results were discussed individually with dancers remotely and where any issues were found from questionnaires, online health tracking or blood tests, timely appropriate tailored recommendations were made.
- Although not the original design, the timing of the study during the COVID-19 pandemic provided insights into the impact of this unprecedented time on dancers. The study showed that easy virtual access to clinical medical support was helpful.
- Dancers and healthcare staff highly rated this initiative of multimodal, personalised monitoring and access to rapid virtual clinical discussion.

*How might this study impact on clinical practice in the future?:*

- Proactive, interactive monitoring of dancer health and wellbeing facilitates personalised, preventative support. It is far better to be proactive in preventing future illness or injury than to deal with issues after they have happened.
- Personalised medical input is important for optimising health and performance and prevention, not just for outcomes injury or illness.
- The dancer health questionnaire could be used for all new dancers entering a dance school or company, to identify any areas where individual dancers might benefit more specific input. For example, to identify any early warning signs of low energy availability in both male and female dancers. In all cases, the objective would be to offer prompt targeted support. The health questionnaire could then be completed annually to act as a monitoring system.
- Comprehensive online monitoring of wellbeing, training load and menstrual cycle for female dancers, is a valuable clinical tool for dancers. Combined with access to individual virtual discussion, this provision is highly rated by dancers and healthcare staff.
- Regular blood testing for dancers provides objective measures of health. This allows early identification of any deficiencies or dysfunction and facilitates prompt advice on appropriate practical measures. This testing can also act as an objective monitoring tool for individual dancers which could be combined with artificial intelligence techniques further personalisation.
- Open access for all dancers to discuss clinical issues with virtual discussions to facilitate ease of access at short notice and on return to performing and touring, which is highly valued by dancers.
- Provision of educational information about relative energy deficiency in sport (RED-S) such as British Association Sport and Exercise Medicine website www.health4performance.co.uk can help the dance community be aware of low energy availability

## Introduction

Lifestyle factors are well established as being important for health. These include behavioural attitudes to exercise, eating a varied diet and having sufficient sleep. Therefore, those with active lifestyles, including professional dancers and athletes, would be expected to be extremely healthy. However, imbalances in behaviours can result in adverse health and performance consequences. Excessive energy expenditure relative to energy intake leads to a situation of low energy availability. Low energy availability is the aetiological factor in the clinical syndrome of relative energy deficiency in sport (RED-S)[1]. RED-S can occur in both male as female exercisers of all levels and ages.

Professional dancers have a demanding schedule from a physical and psychological point of view and unlike athletes, there is no clear off-season of less intensive training and performance. Furthermore, when combined with the aesthetic requirements of dance, this means that dancers can be at risk of low energy availability and developing the adverse clinical and performance outcomes of RED-S.

In the clinical setting, a RED-S diagnosis is confirmed from clinical history, including menstrual status in females, and blood testing to assess endocrine status and exclude medical causes. Endocrine status has been shown to be a good surrogate indicator of energy availability and the outcome of bone stress injuries[2]. These clinical assessments, together with dual-energy X-ray absorptiometry (DXA) scans to assess bone health can be used for risk stratification[3]. However, these metrics only become detectable once the adverse outcomes of low energy availability have occurred. Early detection of exercisers at risk of low energy availability is crucial in order to provide interventions to prevent progression to RED-S.

A recently published dance energy availability questionnaire (DEAQ) identified indicators and correlates of low energy availability and provided a RED-S risk score, allowing it to be used as a screening questionnaire for male and female dancers[4].

The purpose of identifying those at risk of RED-S is to put in place interventions, which include suggested behavioural changes and, in selected individuals, pharmacological treatment to prevent progression and in some cases improve bone health. Evidence for positive outcomes of behavioural interventions is reported in a study of male cyclists[5]. Nevertheless, there is less work on the effectiveness of maintaining energy availability in recovery and the long term[6].

The objectives of this study were to examine the value and effectiveness of health monitoring through a variety of modalities in professional dancers for sustained health and performance.

## Methods

### Study design

A longitudinal study of professional dancers at a dance company to monitor health through a variety of monitoring methods and analyses. The study was approved by the Durham University ethics committee and all participants provided informed consent.

### Recruitment

Professional adult male and female dancers at a dance company were invited to participate from March 2020 through to July 2021

#### Online dance specific health questionnaire

A published dance questionnaire to assess low energy availability was completed by participants to gather medical background, exercise levels, injury history, menstrual status in females, nutrition, subjective report of sleep, fatigue. For each dancer a RED-S risk score was calculated, as previously published[4]. Virtual clinical discussions were conducted by Dr Keay with individual dancers to clarify any areas from the questionnaire.

#### Online monitoring of health and wellbeing

Dancers logged daily wellbeing and training metrics on the monitoring system currently in use at the company. An additional menstrual tracking feature was developed and used by female dancers.

#### Blood biomarkers

Capillary blood markers obtained via finger prick test at standardised time of day, in the morning shortly after waking, were analysed in a UK accredited laboratory to measure thyroid function (TSH), thyroxine (T4) and triiodothyronine (T3), full blood count (FBC), prolactin, follicle stimulating hormone (FSH), luteinising hormone (LH), 9am cortisol, vitamin B12, folate, ferritin, testosterone and oestradiol (for females on day 3 of the menstrual cycle). This occurred at two time points about 6 months apart: when completing the questionnaire and after implementing any advice delivered.

#### Clinical interview

After the initial remote individual interview on completion of the online dancer health questionnaire, regular virtual follow-up occurred to discuss subsequent blood test results, logged wellbeing and menstrual cycle regularity and/or issues. Personalised dance specific actionable advice was provided, with regard to dancer behaviours, including nutrition, recovery/sleep and training load.

#### Feedback

Dancer feedback was sought on the initiatives of multimodal health monitoring and the on demand virtual clinical input.

### Statistical analysis

Data analysis was performed using the open-source tools, SciPy and Pandas (NumFOCUS, Austin, Texas), implemented using the Python programming language. Summary statistics, including count, mean and standard deviation of responses, were calculated for the overall sample and by subgroup, according to the responses to the questions. Body mass index (BMI) was calculated by dividing weight (kg) by the square of height (m). Minimum BMI (BMI min) was calculated based on the dancer’s minimum weight for current height. A weight variability variable was calculated for the current height by dividing the difference between the maximum and minimum weights by the current weight.

## Results

A total of 20 dancers participated in the study (mean age 26.2 years, SD 3.7), comprising 14 females (mean age 25.5 years, SD 3.7) and 6 males (mean age 27.7 years, SD 2.4)

### Energy Availability Questionnaire

The questionnaire results indicated that as a group the participants scored positively. None of the male dancers scored negatively. Negative scores for four female dancers were associated with low minimum BMI (<19), disrupted menstrual function and strong desire to control weight and food.

**Figure 1:**
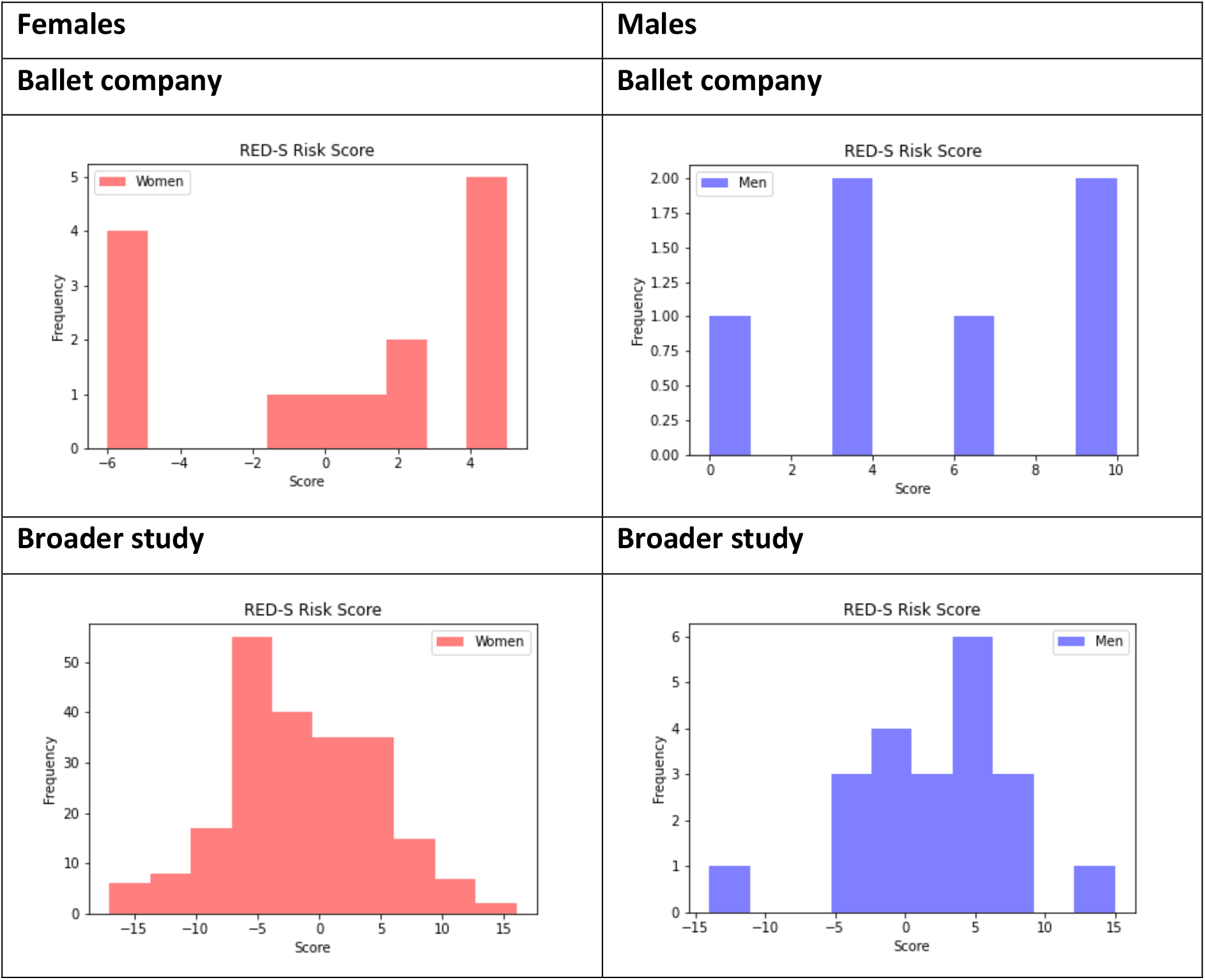
RED-S Risk Scores for company dancers versus previous international dance study

### Hours of training

Table 1 shows that the company dancers complete many hours of exercise per week, with the total ranging from 26 to 60 hours. Even during restrictions to dance studios early in the COVID-19 pandemic, dancers maintained high levels of training with virtual classes and other training modalities.

**Table 1:**
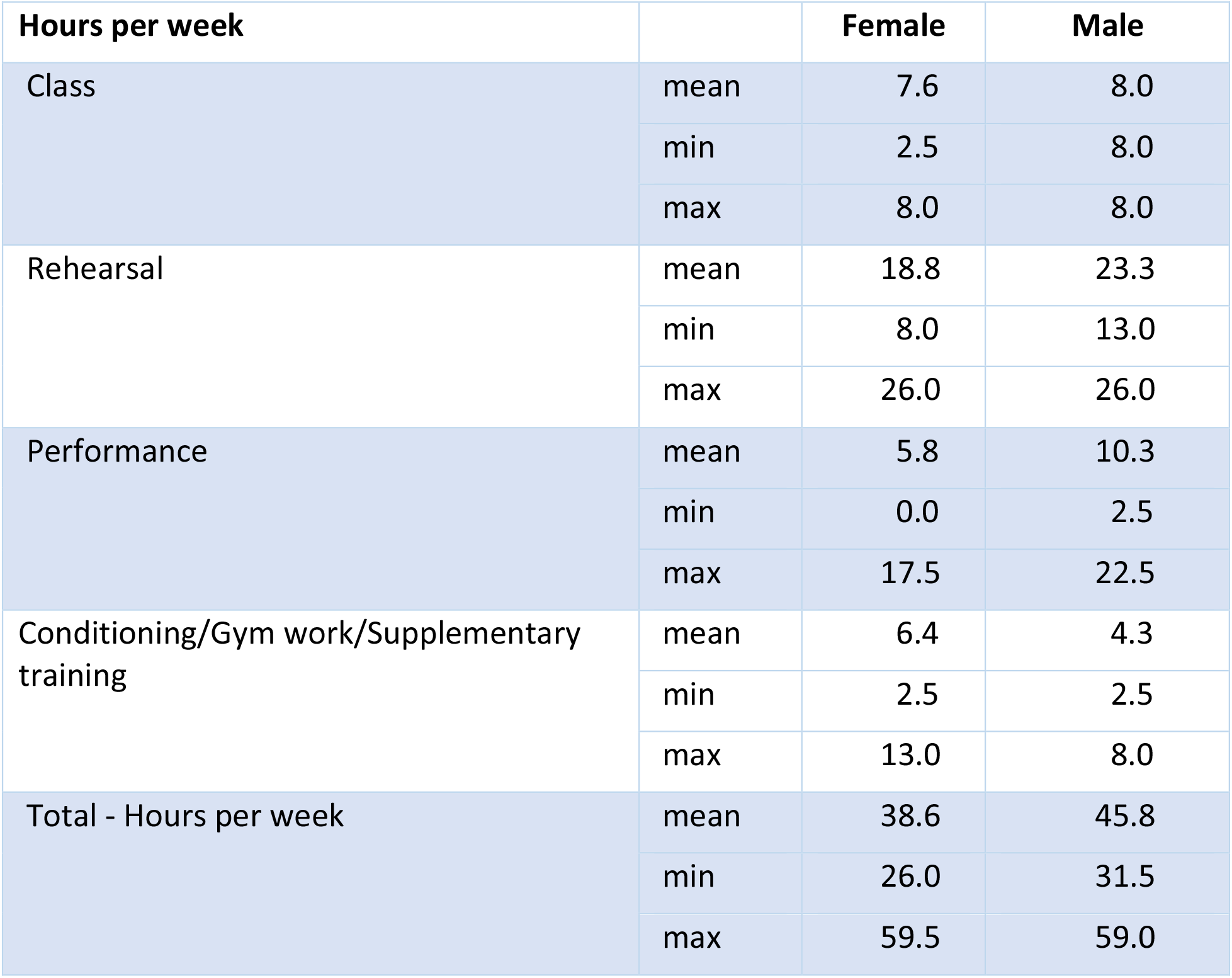
Weekly hours of training

### Anthropomorphic data

Table 2 show the anthropometric data for the dancers indicating that BMI was in healthy range.

**Table 2:** Anthropometric data for dancers

| Anthropometric data |  | Female | Male |
| --- | --- | --- | --- |
| BMI | mean | 19.5 | 22.3 |
|  | std | 1.0 | 1.2 |
|  | min | 18.1 | 21.0 |
|  | max | 21.5 | 24.5 |
| Current height (in metres, e.g. 1.65) | mean | 1.70 | 1.80 |
|  | std | 0.1 | 0.1 |
|  | min | 1.6 | 1.7 |
|  | max | 1.8 | 1.8 |
| Current weight (kg) | mean | 55.3 | 68.5 |
|  | std | 5.0 | 7.7 |
|  | min | 47.0 | 62.0 |
|  | max | 62.0 | 81.0 |

### Wellbeing and psychological report

Dancers were asked to score, on scale of 1 to 5, wellbeing metrics relating to freshness, sleep quality and digestive system function. The results are shown in Table 3. The mean score for both male and female dancers was mid-range.

**Table 3:** Dancer wellbeing scores

| Wellbeing scores |  | Female | Male |
| --- | --- | --- | --- |
| How would you rate your levels of freshness over the past year? | mean | 2.8 | 2.0 |
|  | min | 2.0 | - |
|  | max | 4.0 | 4.0 |
| How would you rate your sleep quality over the past year? | mean | 3.1 | 3.0 |
|  | min | 1.0 | 2.0 |
|  | max | 5.0 | 4.0 |
| How would you rate your digestive system over the past year? | mean | 3.2 | 3.5 |
|  | min | 1.0 | 2.0 |
|  | max | 5.0 | 5.0 |

Regarding psychological factors, the mean scores for both male and female dancers were mid-range for anxiety about missing training. Table 4 shows the scores for rating importance of controlling food intake and weight. The mean score for both male and female dancers indicate a relaxed attitude

**Table 4:** Scores for rating importance of control aspects

| Control scores |  | Female | Male |
| --- | --- | --- | --- |
| Controlling what you eat | mean | 2.7 | 2.7 |
|  | min | 0.0 | 0.0 |
|  | max | 4.0 | 5.0 |
| Controlling what you weight | mean | 2.1 | 1.0 |
|  | min | 0.0 | 0.0 |
|  | max | 5.0 | 3.0 |

### Menstrual function

Twelve of the fourteen female dancers were not on hormonal contraception. Apart from one dancer, these dancers reported eumenorrhoea status at the start of the study. Seven dancers had experienced menstrual disruption in the past during pre-professional training.

### Injury and illness

The dancers had experienced a relatively low level of injury and illness in the past year, with just one or two experiencing longer periods off. This included one female dancer with significant lower leg bone stress injuries and past history of RED-S, before starting her professional career.

### Blood markers

Initial blood testing for the dancers overall indicated that measured markers for full blood count, haematinics and endocrine function were in normal ranges. One female dancer had slightly low iron levels. Two dancers had lower end range vitamin D and were advised about supplementation. One dancer had somewhat high vitamin D associated with high dose supplementation.

For the four females dancers with negative RED-S risk scores, T3 was lower end of range and cortisol upper end of range. Regarding female hormones, one of these dancers had low range female hormones (FSH, LH, oestradiol) and another was on the combined oral contraceptive pill, so it was not possible to comment on her internal female hormones.

Subsequent blood testing indicated that across the group there was a trend to increased levels of “stress” response hormones cortisol and prolactin. Nevertheless, improvement in terms of iron, vitamin D and T3 status was seen in these dancers after receiving personalised feedback and advice on these results.

### Clinical Interviews

At initial virtual individual interview, dancers reported that they felt well-supported by artistic and healthcare staff to deal with the heavy workload and performance schedule. However, at subsequent interview all the dancers reported the challenges during the restrictions of the pandemic, where they were not allowed to enter their dance building for either training or in person clinical support. There was no possibility of performance as theatres were closed. Dancers reported that this was a difficult and stressful situation. Some dances reported symptoms consistent with COVID-19 infection.

The easy access to on-demand virtual clinical discussion was universally reported as being very helpful and eagerness to maintain this approach was expressed from dancers and staff.

### Online monitoring

Dancers logged daily well being showed overall a good level of health. At an early stage some dancers logged symptoms consistent with COVID-19 infection. This was during the initial pandemic lockdown, so dancers were already effectively self-isolating at home. Menstrual cycle tracking was a well-received additional feature and modified in conjunction with female dancer feedback. Tracking showed that although dancers reported eumenorrheic status on the questionnaire, some dancers recorded some menstrual disruption during the lock down during the pandemic from spring 2020.

## Discussion

The results from dancer health questionnaire, monitoring and blood testing showed that the majority of dancers were in good health. These health monitoring initiatives, together with access to on demand virtual clinical discussion were well received by dancers and healthcare staff. The intention is to continue and refine these initiatives beyond the study to maintain optimal dancer health and performance.

### Activity Specific Health Questionnaire

A validated questionnaire exists for identifying low energy availability specifically in female athletes[7]. For male athletes, an activity specific questionnaire, combined with clinical interview, was effective at identifying those with low energy availability and poor bone health[5]. However, there are no validated questionnaires to identify low energy availability and the risk of developing the adverse clinical outcomes of RED-S, which are specific to exercise type and applicable to male and females. A dance-specific energy availability questionnaire demonstrated that male and female dancers can be at risk of developing RED-S[4]. The questionnaire results from the current study, found that only four female dancers out of the whole study group scored negatively. Most likely this is because, in order to be a successful professional dancer and secure employment contracts, it is necessary to be healthy and not susceptible to frequent injuries. For the company in this study, all the dancers had received an educational information discussion about RED-S about 9 months before the start of the study. This could be a contributing factor to the generally healthy disposition of the dancers.

Bone stress injuries are reported as a frequent outcome in athletes with low energy availability, causing endocrine disruption[2]. There is also evidence that bone mineral density may not be recoverable after retirement, as lower than expected for age bone mineral density (BMD) was found in premenopausal retired female dancers[8]. So early identification of dancers at risk of low energy availability with a dance-specific questionnaire, before adverse outcomes of RED-S occur, is a valuable preventative strategy. Combined with clinical interview and objective blood markers, this becomes a very useful, practical clinical tool.

### Blood test results

Initial blood testing showed good correlation to the questionnaire and interview reports. Overall, the dancers were healthy from both subjective report and objective blood measures

Replete vitamin D is recognised as being important for bone, muscle and immune function and in specially in dancers as being a factor in injury reduction[9]. All the dancers in the study were taking vitamin D supplementation, apart from two dancers who were advised to start taking good quality supplementation, especially during lockdown, when time outside at a northern UK latitude was restricted. A female dancer reporting fatigue was found to have slightly low ferritin levels and given appropriate dietary advice. At follow up discussion she reported improved energy levels.

Repeat blood tests across the group showed that these areas of supplementation and nutrition had been addressed. However, the group as a whole recorded increased upper range cortisol and prolactin, in parallel with increased “stress levels” reported at interview, due to not being able to gain access to studios or theatres and without uncertainty about return. This highlights the value of discussing individual findings in the personal clinical context of each dancer.

### Monitoring dancer health and on demand virtual clinical support

Among the female dancers who reported previous menstrual disruption due to dietary restriction during their pre-professional training, one had experienced clinically significant bone stress response issues requiring time off dancing. With appropriate clinical support and monitoring during the study, she returned to full dance activities and received a promotion. For dancers not on hormonal contraception, monitoring menstrual cycles is effectively a non-invasive monthly health check and personalised training metric. This addition to dancer health monitoring received positive feedback from dancers and facilitated remote monitoring and support by clinical staff where indicated. Personalised advice regarding any reported menstrual cycle issues could be provided to ensure dancers felt able to dance to their best throughout their cycle.

### Future applications and feedback

The dance specific questionnaire is a convenient health questionnaire which can be employed in dance schools and companies worldwide. This provides a unified way to assess dancer health and identify dancers who would benefit from personalised input and support from healthcare staff. Menstrual tracking for female dancers together with monitoring blood tests to assess individual response to workload provides objective, quantifiable measures to support optimal dancer health and performance. Combined with easy access, on demand virtual clinical support was found to be effective and well received by dancers. Universally dancers rated this personalised approach with the highest score on the feedback questionnaire.

### Limitations and further work

Since the dancers all came from the same dance company and have received same educational information, selection bias could play a part in these generally positive results. It would be interesting to conduct similar studies in other professional dance companies, across a range of dance genres.

Further validation work of exercise specific questionnaires would be informative. Although there is an assessment tool for risk stratification of those athletes with RED-S[10], this requires input of results from DXA scanning, which might not be accessible or possible from budgetary point of view. Furthermore, this type of scanning to assess bone health utilises ionising radiation, albeit a low dose. Nevertheless, this limits the frequency of repeat scanning to monitor change in bone health. A portable, radiation free method of assessing bone health has shown potential[11].

Further work is focused on the clinical application of artificial intelligence techniques to model female hormones over the menstrual cycle. This approach can identify subclinical anovulatory cycles that occur in low energy availability and can be the precursor to functional hypothalamic amenorrhoea (FHA) if energy deficit becomes chronic. This clinical tool can also be helpful in guiding the restoration of hormone network function in recovery from FHA[12].

## Conclusions

The clinical application of a dance specific health questionnaire screening tool, testing blood markers, combined with comprehensive health monitoring and remote clinical discussion were initiatives successful in identifying dancers who would benefit from personalised advice in the optimisation of health and prevention of RED-S. Dancers rated the opportunity to discuss health and well-being, in confidence, with a doctor conversant in dance with the highest feedback score. In context of multidisciplinary healthcare team, this initiative facilitated personalised advice and support for dancers.

## Data Availability

Data is available on reasonable request

## Statements

## Acknowledgments

Thank you to the company artistic and healthcare staff and all the dancers for their interest and participation in this study.

## Contributorship

NK, ML and GF: conceptualisation of project, development of study design, data collation and analysis, drafting and revision of the manuscript.

## Data sharing

No unpublished data were used in the preparation of this manuscript

## Ethical Approval

This study was reviewed and approved by Durham University research ethics committee.

